# Assessing Rehabilitation Systems in Conflict: A Policy Analysis of Prosthetic and Orthotic Services in Gaza, Palestine

**DOI:** 10.64898/2026.07.01.26357060

**Authors:** Francesca Riccio-Ackerman, Tarek Meah, Khamis Elessi, Haidar Alkhatib, Fatima Mohammadi, Rima Afifi, Hugh Herr, Deema Totah

## Abstract

The Gaza Strip is facing unprecedented levels of destruction to its healthcare system—including the provision of rehabilitation services—since the Israeli Defense Forces began its most recent military operation in October 2023. The Rehabilitation Task Force (RTF) is the governing body tasked with overseeing the provision of these services, including prosthetics and orthotics (P&O), in Gaza. Using the RTF’s published documents—including reports on the state of P&O services, technical reports, and manuals/guidelines for various forms of rehabilitation services—this study aims to assess the capacity of Gaza’s P&O sector to meet the needs of Palestinians seeking P&O services. The study team analyzed the RTF’s documents on P&O capacity and generated ratings according to the World Health Organization’s *Standards for Prosthetics and Orthotics*. The results of our study demonstrate that, despite the extremely challenging circumstances, Gaza’s P&O sector excels in the domain of Policy. The targeted destruction of the rehabilitation sector and intentional withholding of resources preclude the Gaza Ministry of Health from meeting the standards in the domains of Products, Personnel, and Provision of Services. This systemic erosion of rehabilitative medical care translates into a reduced capacity to restore function and mobility to the tens of thousands of people in Gaza currently in need of such care. As such, urgent measures are required to restore, uphold, and expand the capacity of the P&O sector to serve the rights of persons with disabilities in Gaza.

## Introduction

The Gaza Strip, which is part of the internationally recognized occupied Palestinian territory (oPt), has faced unprecedented levels of destruction since the Israeli Defense Forces (IDF) began its most recent military operation in October 2023. In the past two years, at least 78,000 civilians have been killed^1^ and 170,000 have been injured, about 25% of whom need immediate and ongoing rehabilitation services.^2^ Moreover, the military operation has catalyzed the widespread destruction of health services, including essential rehabilitation services.^3^ This “medicide”^4^ translates into a reduced capacity to meet the immediate needs of, as well as to restore function and mobility to, the tens of thousands of people in Gaza currently in need of such care.

This is not the first operation of its kind in Gaza. Since imposing a blockade in 2007, Israel has waged five military operations (2009-2010, 2012, 2014, 2021, 2023-2025) on Gaza^5^, in addition to its militarized response to protesters during the Great March of Return (GMoR)^6–8^. In fact, responding to the challenges facing the rehabilitation sector following the GMoR, local and international actors established the Rehabilitation Task Force (RTF).^9^ The RTF—a group led by the Gaza Ministry of Health (MoH), the World Health Organization (WHO), and Humanity and Inclusion (HI)—is the governing body tasked with overseeing the provision of rehabilitation services, including prosthetics and orthotics (P&O), in Gaza.^9^ Moreover, the RTF periodically produces reports on the state of P&O services, technical reports, and manuals/guidelines for various forms of rehabilitation services.

Using these sources, the current study aims to assess the capacity of Gaza’s rehabilitation system—particularly P&O care—to meet the needs of Palestinians with disabilities. We begin by providing a background on P&O care in Gaza. Next, we describe our method for collecting and assessing data from the RTF’s public-facing documents. Then, we present our results before discussing how and why the ongoing Israeli assault remains the greatest challenge to the effective delivery of P&O services in Gaza. Finally, we conclude by outlining practical recommendations for improving P&O assessment to inform capacity strengthening efforts in active crisis settings.

## Methods

### Conceptual Frameworks

Two frameworks guided our analysis:

### The World Health Organization (WHO)’s *Standards for Prosthetics and Orthotics*

These standards provide guidance to UN Member States to consider and implement in the establishment and maintenance of “appropriate systems and infrastructure for the provision of high-quality prosthetics and orthotics services.”^10^ Published in 2017, the two-part document is a collaboration between the WHO, the US Agency for International Development (USAID), and the International Society for Prosthetics and Orthotics (ISPO).

The document is divided into two parts: “Part 1: Standards”^10^ and “Part 2: Implementation Manual.”^11^ One of its central aims is to “ensure that prosthetics and orthotics services are people-centered and responsive to every individual’s personal and environmental needs.”^10^ The framework includes four core areas—policy, products, personnel, and provision of services (the “4 P’s”):

1. **Policy** - emphasizes that governments are responsible for ensuring affordable, accessible, and high-quality P&O services through strong policy frameworks, and (ideally) universal health coverage.
2. **Products** - highlights the need for P&O products to be appropriate to the local context and affordable, supported by a national priority list to guide procurement and resource allocation.
3. **Personnel** - emphasizes the need for adequate training, regulation, and professional recognition of P&O personnel to ensure service quality and protect users, and highlights the importance of a multidisciplinary team approach, particularly for individuals with complex physical impairments.
4. **Provision of Services** - advocates for user-centered P&O services, where individuals are empowered to make informed decisions and actively participate in all aspects of care and service development.

### The Convention on the rights of Persons with Disability

On December 12, 2006, the United Nations General Assembly adopted resolution A/RES/61/106, establishing the *Convention on the Rights of Persons with Disabilities* (CRPD). The CRPD obligates State Parties to “promote, protect, and ensure the full and equal enjoyment of all human rights and fundamental freedoms by all persons with disabilities”^12^ through the provision of certain guarantees and liberties. The CRPD also applies to persons with disabilities in the territories over which a ratifying State Party “exercises jurisdiction” or has “effective control.”^13^ Israel signed the treaty in 2007 and ratified it in 2012, thereby assuming legal obligations towards persons with disabilities, including Palestinians living in the oPt. In 2014, the State of Palestine (as a “nonmember observer” of the UN) also acceded to the Convention.

Given the 4 P’s alignment with core obligations under the CRPD, in addition to its relevance to rehabilitative health systems in State Parties, we focus our analysis on the standards within the framework.

### Data Collection

The study team analyzed the RTF’s public-facing documents on P&O care and capacity and generated ratings according to the 4 P’s framework.^10,11^ As of August 2025, the RTF has published eight reports on their website. These documents represent the most authoritative and contemporaneous accounts of P&O in the Gaza Strip. Since all documents were published between October 2023 (the start of our study period) and August 2025, they were all considered for inclusion into the study. **Error! Reference source not found.** notes which were included or excluded and the corresponding rationale.

**Table 1.** Status of RTF documents -included or excluded and rationale for exclusion.

|  | Report Type | Included? If not:<br>explain why. | Report Title | Publication Date |
| --- | --- | --- | --- | --- |
| 1 | Situation Report | Yes | Rehabilitation SitRep Q2 2025 | June 11, 2025 |
| 2 | Manual and Guideline | Yes | Minimum Rehabilitation Service Package for Gaza, oPt - Based on WHO Standards | May 5, 2025 |
| 3 | Situation Report | Yes | Rehabilitation Task Force SitRep February 2025 | May 3, 2025 |
| 4 | News and Press Release | Yes | WHO analysis highlights vast unmet rehabilitation needs in Gaza | September 12, 2024 |
| 5 | News and Press Release | No; Document does not provide information about Gaza. | Ministry of Health Inaugurates First Rehabilitation Outpatient Centers in West Bank with Funding from the Republic of Korea and Support from WHO and UNOPS | May 6, 2025 |
| 6 | Assessment | No; Document does not include information specific to P&O policy, service delivery, or systems of provision. | Estimating Trauma Rehabilitation Needs in Gaza Using Injury Data from Emergency Medical Teams (30 July 2024) | September 12, 2024 |
| 7 | Manual and Guideline | No; Document does not include information specific to P&O policy, service delivery, or systems of provision. | Rehabilitation Task Force Technical Note: Spinal Cord Injury Care in Gaza | May 6, 2024 |
| 8 | Manual and Guideline | No; Document does not include information specific to P&O policy, service delivery, or systems of provision. | Rehabilitation Task Force Technical Note: Amputations and Prosthetics in Gaza | May 6, 2024 |

### Data Analysis

To assess the degree to which the P&O services in Gaza were meeting the specific WHO standards, the study team devised a four-point categorical rating scale:

- Unknown: cannot make an informed decision given that specific information about the standard is not available in the evaluated data sources;
- 1: the standard was identified in the document but was not met to any extent;
- 2: the standard was identified as a goal and partially achieved; and
- 3: the standard was identified as a goal and fully achieved.

Two members of the research team (TM, FRA) independently and separately reviewed the documents listed in **Error! Reference source not found.**, recording preliminary ratings for each WHO standard. Following independent review, the two raters convened to reconcile discrepancies and to develop a preliminary consensus. This was subsequently circulated to two additional team members (RA, DT) both with expertise in rehabilitation and public health systems, for critical review, comments, and questions. Final ratings were then determined through discussion to achieve consensus among five team members (TM, FRA, RA, DT, FM). *Data Organization*

The WHO Standards for P&O are organized into a hierarchy (see Fig 1), beginning with the 4 P areas. In the original WHO document, some but not all areas cascade down to several major categories (e.g. Policy is divided into “Leadership and Governance” and “Financing”).

**Fig 1.**
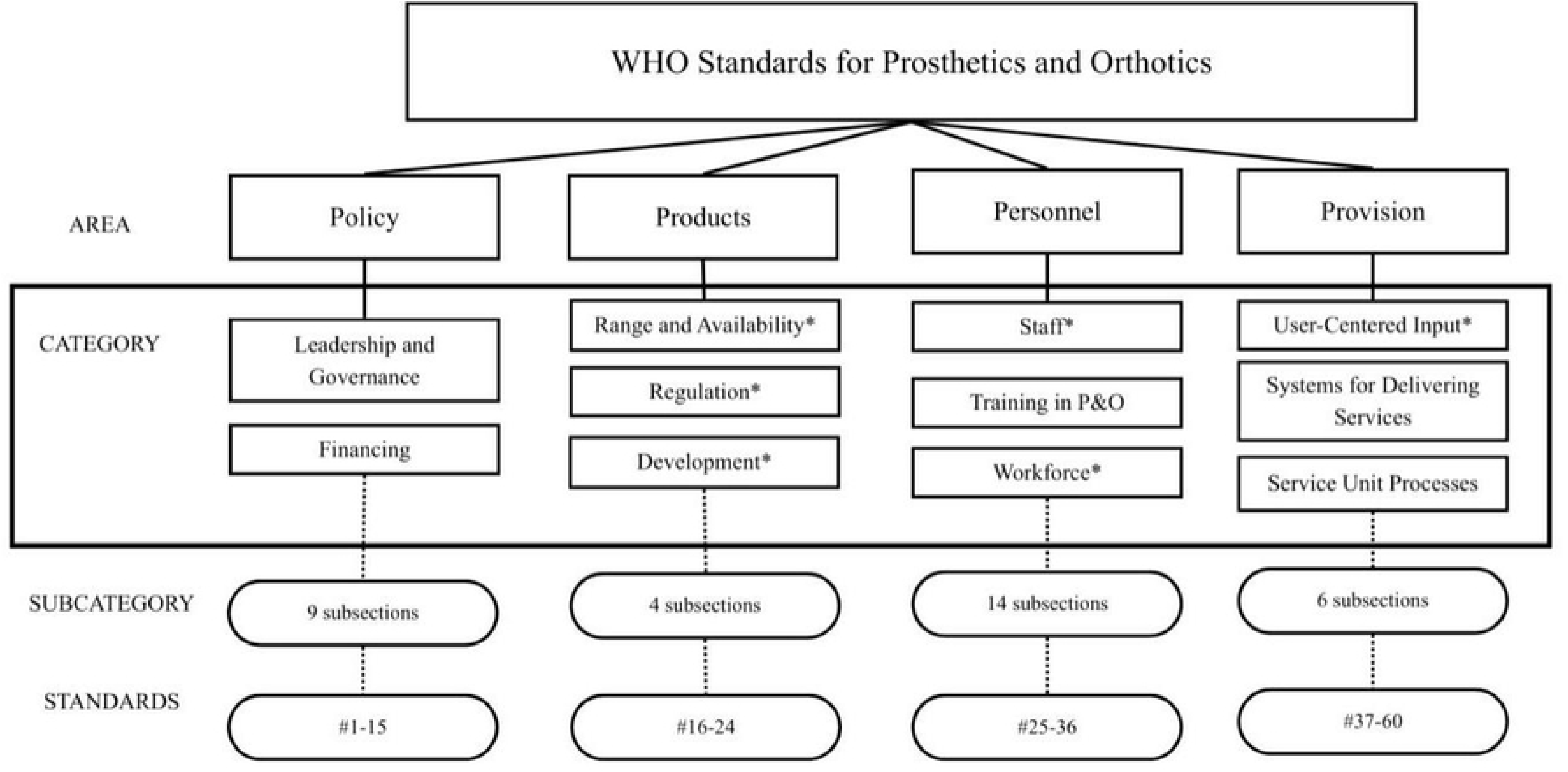
WHO Standards for Prosthetics and Orthotics

Each category is then separated into Subcategories (e.g., “Leadership and Governance” consists of “Stakeholders and Coordination,” “Guiding Framework for Provision,” “Monitoring,” etc. and each subcategory includes at least one standard (e.g., “Stakeholders and Coordination” includes three individual standards. Several subcategories were not couched within a larger category and were given a category title by the study team; these are marked with an asterisk in Fig 1. There were sixty total P&O standards across four areas.

The study team assessed and provided ratings for all sixty standards. For simplicity, we present results at the subcategory level. For complete detailed ratings at category, subcategory, and individual standard levels, refer to S1 Table. The study team assessed and provided ratings for all sixty standards. However, we present results at the level of the category only. Complete detailed ratings at the category, subcategory, and individual standard level are available in S1 Table for reference.

### Ethics Statement

This study did not require an ethics review since it analyzed publicly available secondary data and did not include human participants.

### Positionality Statement

The authors of this paper are Palestinian, have Palestinian relatives, and/or are academics in engineering or the social sciences. Several have firsthand experience of Israeli aggression in Palestine (West Bank and Gaza) or Lebanon. One author is a person who has experienced the loss of two limbs, has lived experience with this disability and has access to advanced P&O care and assistive technology/devices. Another author has an immediate family member with an amputation. We sought to mitigate author bias by relying on WHO reports and frameworks for analysis. We also sought to mitigate power imbalances by including research from Palestine, specifically Gazan health care sources. The authors include professionals in healthcare living in Gaza at the time of writing.

## Results

Category ratings indicate current strengths and challenges of the Gaza’s P&O sector. We summarize details for each category below. **Error! Reference source not found.** provides the text for each standard under the category/subcategory, and S1 Table includes standard-level ratings.

**Table 2.**
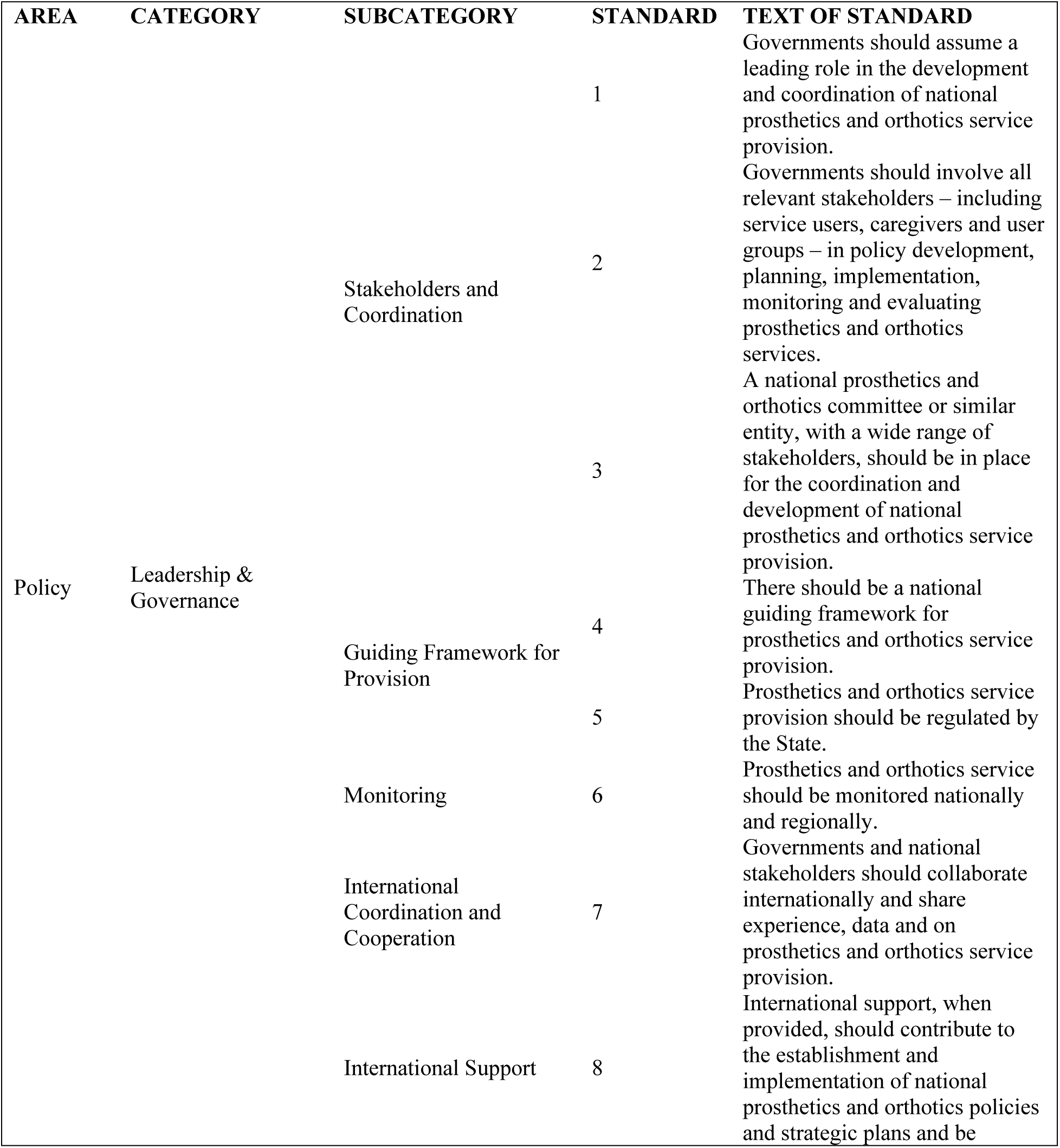
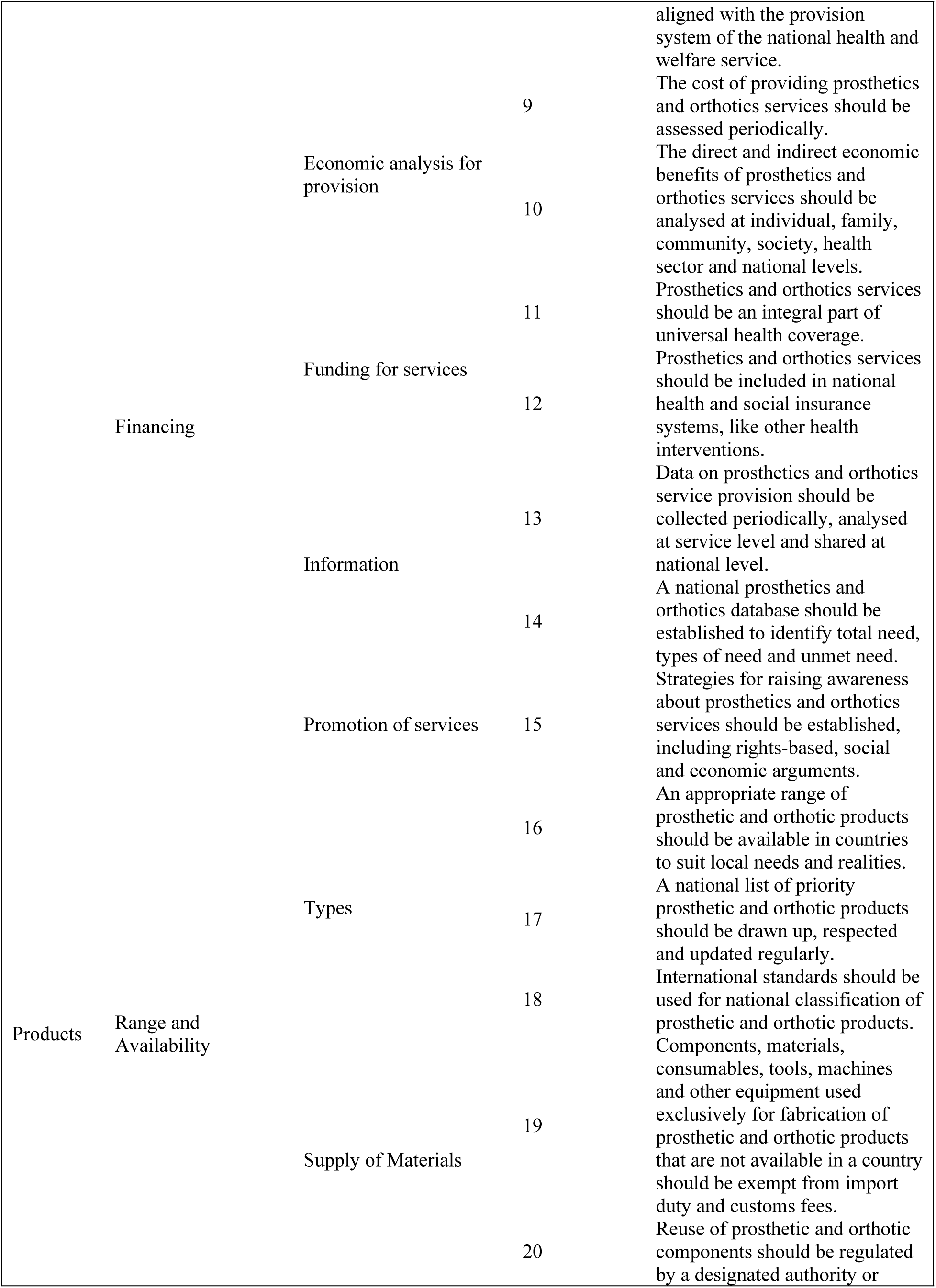
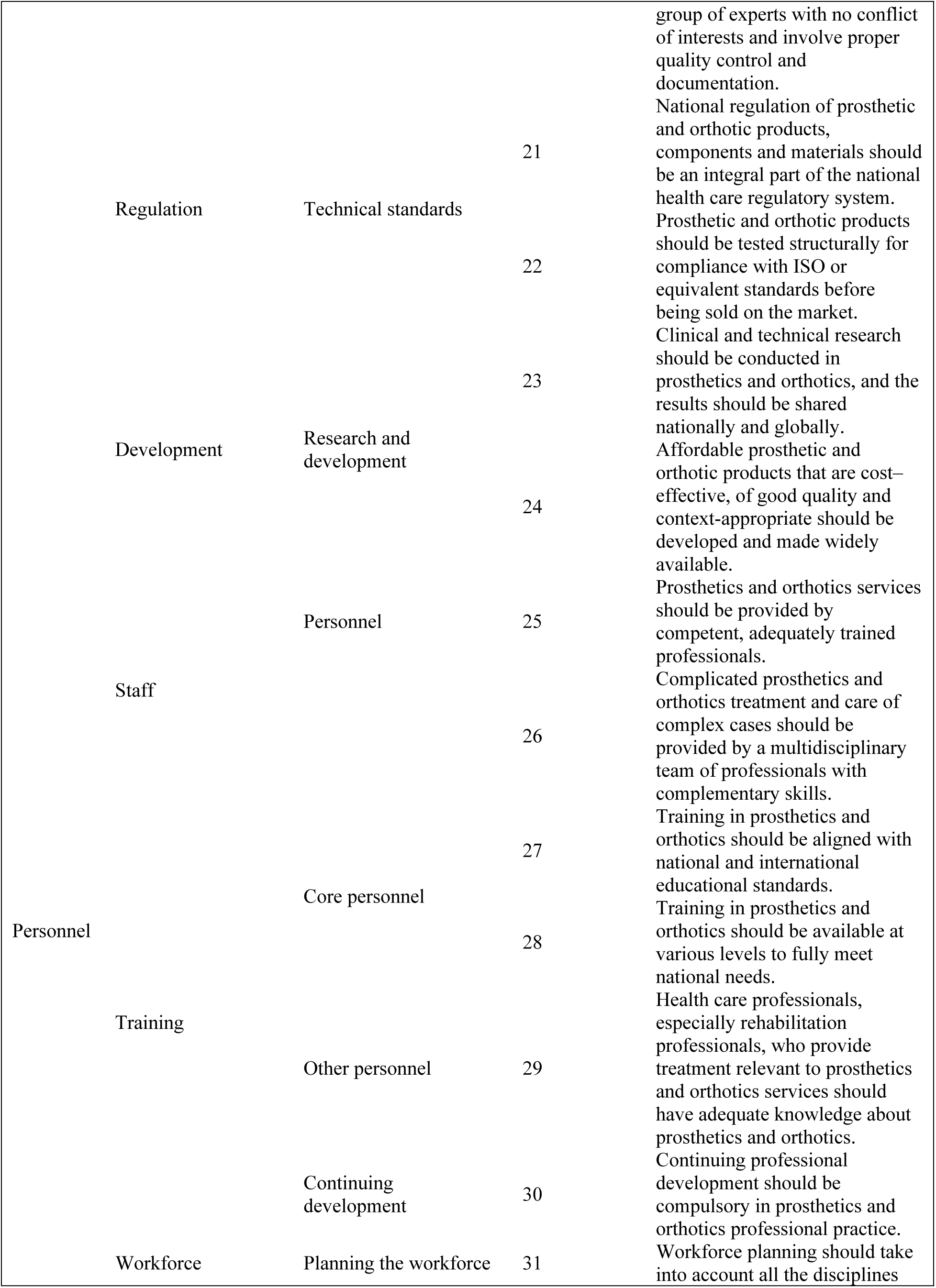
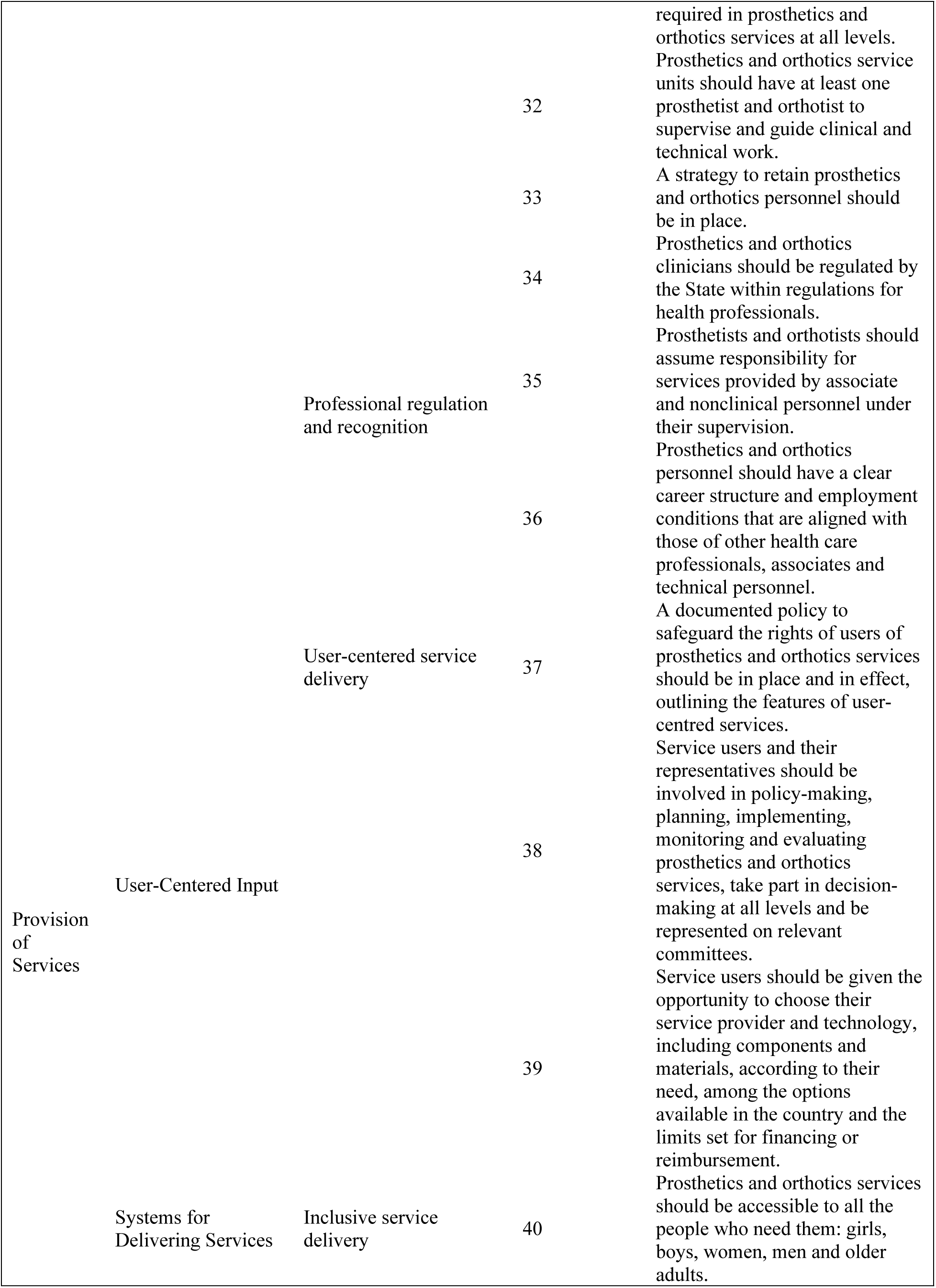
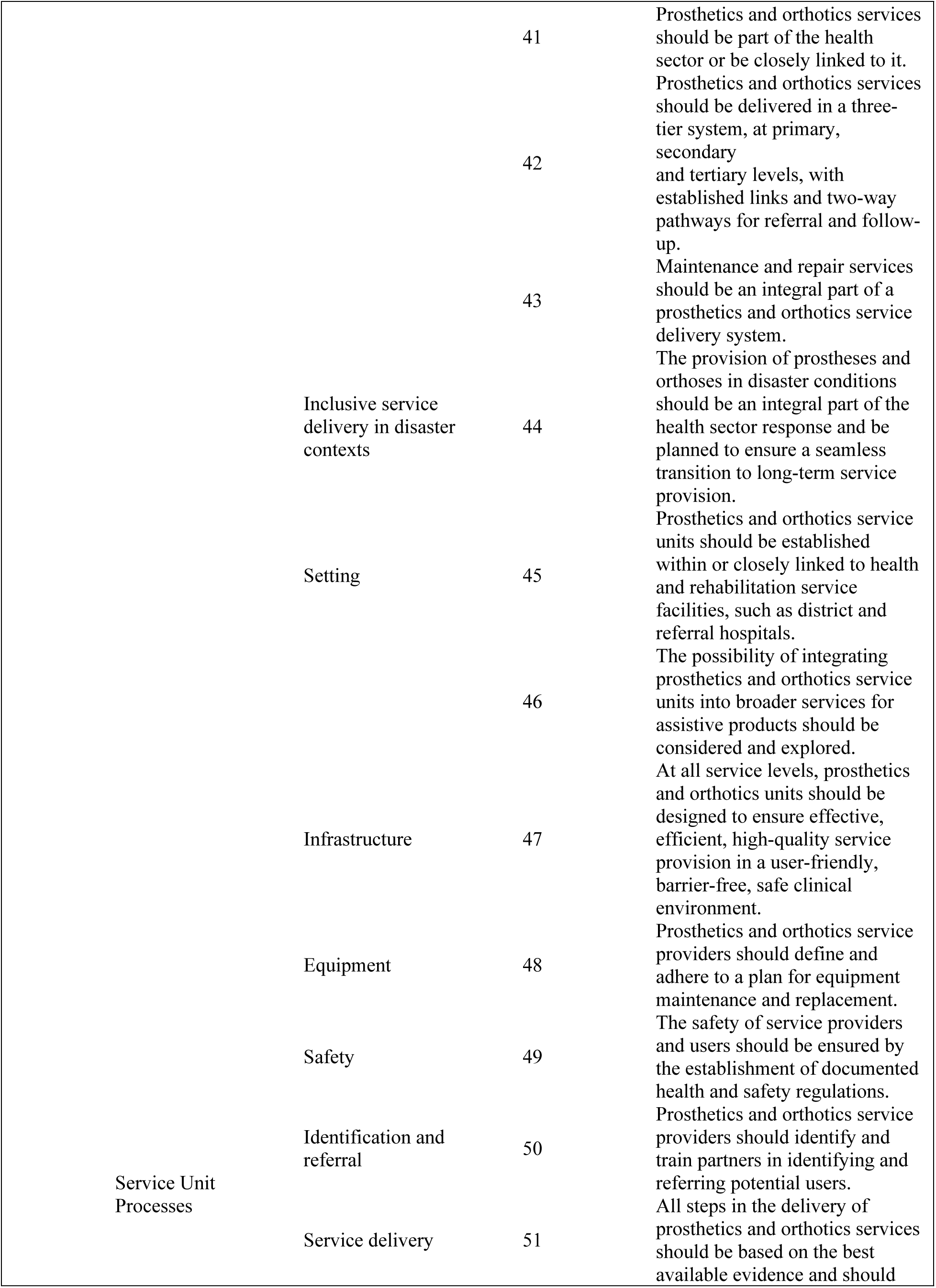
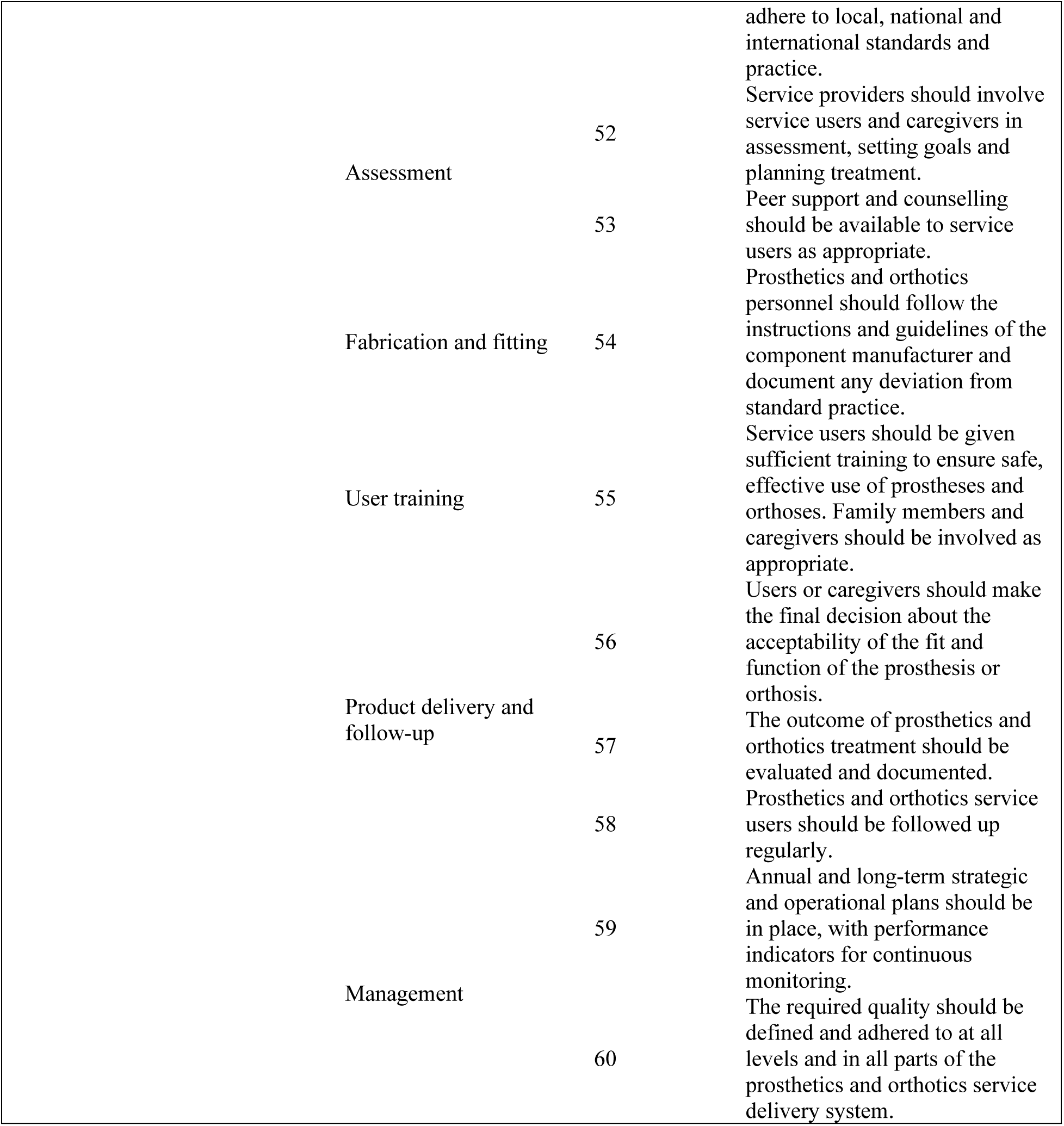
WHO Standards for Prosthetics and Orthotics, Part I.

The overall category scores demonstrate that, despite the extremely challenging circumstances, Gaza’s P&O sector excels in the domain of Policy. However, the targeted destruction of the rehabilitation sector^14^ and intentional withholding of resources^15^ preclude the Gaza MoH from meeting the standards in the domains of Products, Personnel, and Provision of Services (Fig 2).

**Fig 2.**
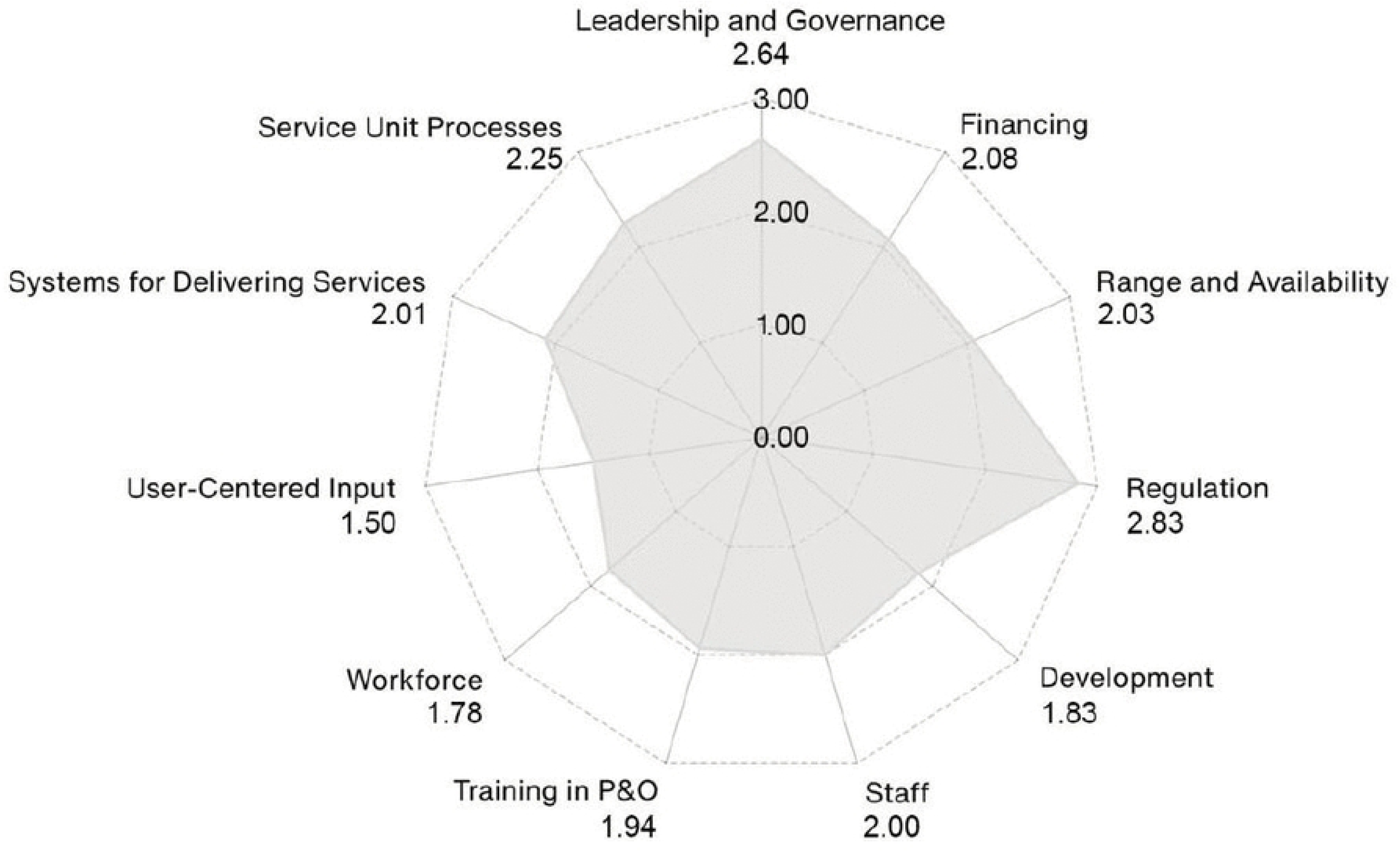
Category-level ratings produced from averaging the subcategory ratings. Individual standard ratings within each subcategory are in S1 Table.

### Policy

#### 1. Leadership and Governance – 2.64/3.00

This score is reflective of the RTF’s commitment and success in guiding and monitoring the provision of formal P&O services in Gaza. This process requires significant international cooperation, as demonstrated by WHO’s and H&I’s participation in coordinating and supporting on-the-ground interventions. However, while the RTF specifies alignment with WHO guidelines for P&O care and ISO standards for the development and provision of P&O services,^2,3^ there is little evidence that suggests the body is regulating and/or monitoring the provision of informal services.

#### 2. Financing – 2.08/3.00

The RTF excels in providing costs and raising awareness about P&O services, yet, to the best of the study team’s knowledge, does not provide economic analyses of the direct and indirect benefits of these services in public-facing reports. Furthermore, from the RTF’s reports, it is unclear whether P&O services are considered part of universal health coverage.

### Products

#### 1. Range and Availability – 2.03/3.00

The RTF provides a list of priority P&O products and abides by international standards to classify these needs. However, the RTF reports that the lack of equipment precludes service delivery in 100% of inpatient rehabilitation services, 100% of prosthetic and orthotic services, and 88% of outpatient services.^3^

#### 2. Regulation – 2.83/3.00

The P&O products which the RTF regulates are an integral component of the health care regulatory system in Gaza. Additionally, these products—because they are mostly provided by international NGOs—already meet ISO or equivalent standards. Moreover, the MoH has committed to upholding this compliance, per the Minimum Rehabilitation Service Package (MRSP).^16^

#### 3. Development – 1.83/3.00

According to the MRSP, the MoH has stated a commitment to developing affordable, high-quality, and context-appropriate products.^16^ However, the ongoing military hostilities and siege of medical supplies have resulted in (per the February and June situation reports) notable gaps in assistive products for people with existing injuries^3^ and insufficient materials^2^ to address all the needs.

### Personnel

#### 1. Staff – 2.00/3.00

Appropriate training is available for existing staff, however capacity is extremely limited^3^, especially in light of the massive increase of individuals requiring P&O services. Furthermore, Israel’s targeted killings and forced displacement of healthcare workers has further reduced the P&O workforce.^17^

#### 2. Training in P&O – 1.94/3.00

Given the limited number of P&O personnel operating in Gaza and the ongoing crisis, the RTF has invested its resources in meeting the immediate needs of patients with disabilities, as opposed to longer-term training. Despite the challenges, however, the RTF is assisting in the development of a national training framework for rehabilitation with its partners.^2^ UK-Med, for example, has been preparing a capacity building initiative via online training as of June 2025.^2^

#### 3. Workforce – 1.78/3.00

Current conditions severely restrict meaningful workforce planning as healthcare professionals have been forced to flee or have been detained and/or killed. Moreover, per the RTF’s June 2025 situation report, there were only 9 trained prosthetists and orthotists (CPOs) in Gaza,^2^ serving a population of 2.1 million people.^18^ However, per the WHO standards—which recommend 5-10 P&O clinicians per million population—there should be between 10 to 20 CPOs.^12^

### Provision of Services

#### 1. User-Centered Input – 1.50/3.00

To the best of the study team’s knowledge, the RTF has not reported on user-centered input in its public-facing documents.

#### 2. Systems for Delivering Services – 2.01/3.00

Challenges to delivering services include destruction of facilities, lack of equipment and supplies, and staff shortages.^3,17^ Despite this, however, the RTF is able to maintain close linkage with the health sector and coordinate service delivery across primary, secondary, and tertiary levels, ensuring established referral pathways for individuals requiring prosthetic and orthotic care.^16^

#### 3. Service Unit Processes – 2.25/3.00

Per the RTF reports, multi-disciplinary rehabilitation services exist throughout the Strip, and a referral network is described.^2,3^ Moreover, data and outcomes are collected and shared at multiple health delivery service unit (HDSU) levels for rehabilitation services in the WHO’s Health Resources and Services Availability Monitoring System.^3^ However, the limitation in this category, per the RTF, is meeting the volume of services required and following up with treated patients because of continuous forced displacement.^3^

## Discussion

This paper assessed the post-October 2023 state of rehabilitation services—specifically P&O care—in Gaza, oPt using the WHO’s 4 P’s framework. Our analysis enables a structured evaluation of both systemic strengths and opportunities for strengthening rehabilitative care. A contemporaneous understanding of the P&O sector is critical to measure the damage against persons with disabilities, to assess the services needed to ensure their rights and wellbeing, and to inform rebuilding efforts. The findings demonstrate that, despite the existence of policy frameworks, the systematic destruction of the healthcare system—including infrastructure, workforce, and resources—has severely limited Gaza’s capacity to provide meaningful care; this is despite RTF members’ demonstrated awareness of international standards, earnest attempts at meeting them at all levels, and their commitment to documentation. This systemic erosion of rehabilitative medical care translates into a reduced capacity to restore function and mobility to the tens of thousands of people in Gaza currently in need of such care.

### Israeli Violations of the CRPD

Israel’s current military actions in the oPt have raised serious concerns regarding non-compliance with the provisions of the CRPD^12^ and these concerns have intensified over the past 23 months. Israeli forces have destroyed rehabilitation warehouses and facilities,^4,14^ severely restricted access to medical supplies,^15^ targeted medical staff,^19^ and displaced populations with disabilities,^20^ resulting in conditions that undermine the rights guaranteed under the CRPD including, but not limited to, freedom from violence (Article 16), liberty of movement (Article 18), personal mobility (Article 20), and access to habilitation and rehabilitation (Article 26).

According to the United Nations Relief and Works Agency (UNRWA), persons with disabilities in Gaza currently face “disproportionate risks and challenges,” including physical and informational barriers, as well as electricity-depleted assistive devices.^21^ Additionally, the UN Committee on the Rights of Persons with Disabilities noted that persons with disabilities in Gaza fear being “the first and the next to be killed,”^20^ due to their limited ability to flee. Such infringements reveal not only systemic collapse in Gaza’s health sector, but also Israel’s violation of its own commitment to the CRPD, and its ongoing obstruction of the State of Palestine’s ability to meet its obligations under the CRPD. These infringements on the rights of Palestinians with disabilities—as guaranteed to them by the CRPD—are not new. Scholars have long documented how Israeli military campaigns in Gaza have strained the already fragile healthcare system in Gaza.^22,23^

### Limitations of the Current Study

The first limitation to the current study is data availability. As a first attempt to assess the quality of the P&O sector in Gaza, this analysis utilized only publicly available RTF documents in English, as official communication from the leadership in the rehabilitation sector of Gaza.

Direct fieldwork with and first-hand accounts from patients, healthcare workers, and health sector leaders would greatly contribute to a more comprehensive understanding of rehabilitation services in Gaza.

Another key limitation to consider is the scope of existing WHO P&O standards. These standards were not specifically developed for use in conflict, developing or destroyed health systems, especially in active crisis settings; thus providing insufficient implementation guidance for developing local P&O capacity in these contexts. The standards also suggest singular pathways for achieving key metrics, as opposed to adaptive strategies to account for resource-limited P&O sectors. Finally, the standards do not thoroughly account for cultural considerations that affect P&O use.

### Implications for research, practice, and action

As noted above, the WHO 4 P’s framework is insufficient for assessing systems in conflict zones, particularly those that are actively targeted and besieged by external forces. Future work should expand upon the existing assessment frameworks to allow identification of the root causes preventing successful fulfillment of rehabilitation services. Such holistic approaches would direct accountability where it belongs—toward the actors and conditions that deliberately undermine health systems—and to call out these violations with evidence, clarity, and moral force.

In response to the ongoing crisis in Gaza, the study team presents a tiered list of critical action items for ensuring P&O rehabilitative medical care that conforms to the WHO 4Ps is available for persons with Disabilities in Gaza (**Error! Reference source not found.**), noting its time periodicity and responsible parties. Given the impact of Israeli military aggression, the most critical action is a permanent end to the Siege and an immediate and permanent cease fire, which also allows Palestinian self-determination. Other immediate, short-, medium-, and long-term actions are recommended.

**Table 3.** Critical Action Items for ensuring minimum P&O rehabilitative medical care is available for persons with disabilities in Gaza.

| Time Period | Actions | Responsible Parties |
| --- | --- | --- |
| *Immediately* | A permanent end to the widespread aggression against the civilian population and infrastructure of Gaza | State of Israel; International Governing and Collaborating Parties |
|  | • End the use of cluster munitions and high velocity weaponry, stop all harm against civilians and especially persons with disabilities<br>• Stop targeted attacks against healthcare workers, first responders, and healthcare facilities<br>• End the siege which indiscriminately causes preventable suffering, disease and death<br>• Provide sufficient food, medicine and humanitarian aid to the population in Gaza, and targeted aid to the injured population<br>• Coordinated entry of conflict medicine specialists, healthcare workers and administrators to assess and assist with immediate rehabilitation needs |  |
| *Short-Term* | • Use of mobile and field-based P&O services<br>• Collaborative assessments of immediate needs and challenges of persons with disabilities and those requiring long-term rehabilitative care | Gaza Ministry of Health; financial supporters; humanitarian organizations |
| *Medium-Term* | • Investment in workforce training (including remote/distance learning models for P&O)<br>• Standardized protocols for triage and follow-up<br>• Rehabilitation integration with trauma care | Gaza Ministry of Health; financial supporters; humanitarian organizations; academic institutions (local and international) |
| *Long-Term* | • Incorporation of P&O into national health and insurance systems<br>• Sustainable financing mechanisms<br>• Stability in the legal and policy frameworks necessary to ensure compliance with international guidelines | State of Israel, Gaza Ministry of Health; financial supporters |

## Conclusion

The findings of this analysis demonstrate that, despite the existence of policy frameworks, the systematic destruction of the healthcare system—including infrastructure, workforce, and resources—has severely limited Gaza’s capacity to provide meaningful care. In line with international obligations, urgent measures are required to restore, uphold, and expand the capacity of the P&O sector to serve the rights of persons with disabilities in Gaza. We urge attention to this critical gap.

## Data Availability

This study used publicly-available data, published by the Rehabilitation Task Force at ReliefWeb Response.

https://response.reliefweb.int/palestine/rehabilitation-task-force

## Supporting Information

S1 Table. WHO Standards.

## Acknowledgments

The authors acknowledge the clinicians and humanitarian partners working in Gaza who, despite the ongoing bombardment, remain committed to providing care to the thousands of civilians who continue to sustain injuries. We also acknowledge the members of the Rehabilitation Task Force, whose data made this analysis possible.

